# Commissioning of Tier 3 obesity services by Integrated Care Boards in England: an analysis of responses to Freedom of Information requests

**DOI:** 10.1101/2024.12.20.24319422

**Authors:** Nicholas Finer, Nikolaos Fragkas, Alexander Dimitri Miras, Sarah Le Brocq, Dimitri J Pournaras, John Wass, Cecilia Pyper

**Affiliations:** Public Health Action Support Team CIC (PHAST), London, UK; Medical Affairs, Pfizer Ltd, Walton Oaks, UK; School of Medicine, Ulster University, UK; All About Obesity CIC, UK; Department of Bariatric and Metabolic Surgery, North Bristol NHS Trust, Southmead Hospital, Bristol, UK; University of Oxford - Oxford Centre for Diabetes, Endocrinology & Metabolism (OCDEM) Churchill Hospital, UK

## Abstract

**Objectives:** This research surveyed Tier 3 adult weight management services in England commissioned by integrated care boards (ICBs) across England in financial year 2022-2023.

**Design:** Survey of public health services commissioned by ICBs gathered via freedom of information requests

**Setting and Participants:** All 42 ICBs in England

**Primary and secondary outcome measures:** The primary outcome measure was whether or not there was Tier 3 provision. Secondary outcome measures included the type of Tier 3 services provided and the estimated number of patients referred and treated.

**Results:** We received responses from all 42 ICBs, a 100% response rate. Five reported that their ICB had no Tier 3 provision (NHS NW London, NC London, Lancashire and South Cumbria, Northeast and North Cumbria, and Northamptonshire).

Deprivation is the major underlying inequality underpinning differences in obesity levels (24, 25), so we explored the relationship with deprivation measured by the Index of Multiple Deprivation (IMD) for each ICB and referral rates to Tier 3 services. Including only data from those ICBs returning numbers of referrals identified there was no correlation between IMD and referral rates (R^2^ = 0.074; P=0.210).

Since obesity prevalence data mapped to ICB level was not available we used regional data and estimated that 25% of people with obesity were likely to have a BMI≥35 kg/m^2^ and be eligible for referral to Tier 3 management. We then calculated the percentage receiving treatment. According to ICN reports the percentage of eligible patients treated ranged from 0 to 1.1%, the highest number was in the South East of England, a region with the lowest levels of deprivation.

Lastly, we noted that, several ICBs were commissioning services that appear to not meet the definition of a Tier 3 service and were more in keeping with the criteria for a Tier 2 service. Accessing full details for these programmes is currently not readily available. However, treatment programmes exclusively online may fail to: be complete in their multidisciplinary team; provide clinical assessment and screening for obesity related diseases; include access to pharmacotherapy or surgery or comply with NICE guidance.

**Conclusions:** Provision of Tier 3 services in England is inadequate and variable and they currently fail to meet the needs of the population. Five ICBs provide no Tier 3 services at all. Areas with highest levels of deprivation provided the most limited access. Even where commissioned, services often do not meet commissioning guidelines. Action is required to implement Health Service Policy and to ensure obesity services conform with clinical need and national guidelines.

**Strengths and Limitations of this study:**

- The study covered all ICBs in England giving an accurate overview of Tier 3 services in financial year 2022-2023.
- The findings rely on the accuracy of the data supplied by the ICBs, which could not be verified.
- Relevant measures of population health including ethnicity, the prevalence of obesity across England and areas of deprivation were used to assess potential need for AWM services across England.
- The study took into account services commissioned from private Tier 3 service providers.
- The period covered was soon after ICBs took responsibility for commissioning Tier 3 services, and in many cases had yet to appoint an ‘obesity lead’, despite their roles and responsibilities being laid out in May 2022.
- In October 2023 and March 2024, NICE approved 7 digital services for prescribing and monitoring and delivering multidisciplinary weight-management services. These value assessments post-dated our survey dates and were not aligned to the definition of Tier 3 services.

## Introduction

Obesity is a complex and multifaceted chronic disease of serious public health concern in England; it is a leading cause of preventable illness and premature death. Understanding the prevalence and needs of this population is essential to ensure the increasingly effective and safe weight loss interventions can be appropriately scaled and targeted to provide appropriate care to people living with obesity. Data from the 2021 Health Survey for England found 26% of adult classified as ‘obese’ and a further 44% classified as ‘overweight’.(1) Considerable geographical heterogeneity in obesity prevalence exists, the North-South divide particularly evident, with obesity rates generally higher in the North of England compared to the South.(2) In the North of England, cities such as Liverpool, Manchester, and Newcastle have been identified as obesity hotspots, with prevalence rates well above the national average. These areas often experience higher levels of deprivation, a known risk factor for obesity. Conversely, the South of England tends to have lower obesity rates, although disparities still exist within this region. Ethnicity is another important determinant of obesity prevalence, with certain minority groups, such as Black, Asian, and Minority Ethnic (BAME) communities, experiencing disproportionately higher rates compared to the White British population.

Obesity treatment in England has been delivered using a tiered approach according to the severity and complexity of the disease. (3–5) Tier 1 and 2 services are defined as public health measures and ‘lifestyle weight management services’ that are time-limited usually to 12 weeks. Both are funded by local authorities. Tier 4 services include bariatric surgery and specialist medical treatment. Tier 3 services are clinician-led, multidisciplinary interventions designed to support individuals who have not responded to conventional weight management interventions and who are at high risk of, or have already developed, obesity-related complications. NHS commissioning guidance has been developed describing the following five components of a tier 3 services:(3)

### Multidisciplinary Approach

Tier 3 services typically involve a multidisciplinary team of healthcare professionals, including a physician, dietitian, specialist nurse, clinical psychologist, liaison psychiatrist, exercise physiologist and physical therapist to provide comprehensive assessment, personalised treatment plans, and ongoing support to address the complex physical, psychological, and behavioural aspects of obesity. Where Tier 3 services are integrated into a Tier 4 surgical service a bariatric surgeon will form part of the team.

### Comprehensive Assessment

Individuals referred to Tier 3 services should undergo a comprehensive assessment to evaluate their medical history, current health status, psychological well-being, dietary habits, physical activity levels, and readiness for change, as well as screening for undiagnosed obesity complications (e.g. type 2 diabetes, obstructive sleep apnoea, metabolic-associated fatty liver disease).

### Personalised Treatment Plans

Based on the assessment findings, personalised treatment plans should be developed in collaboration with the individual, considering their preferences, goals, and medical needs.

### Ongoing Support and Monitoring

Tier 3 services should provide ongoing support and monitoring to individuals throughout their weight management journey.

### Evaluation and Quality Improvement

Continuous evaluation and quality improvement efforts that include feedback from patients and audit are integral to the development of Tier 3 services. While there is extensive evidence on the efficacy (and safety) of the individual components comprising a Tier 3 service, evaluation of Tier 3 services themselves is scant, in part because many Tier 3 services may either not be ‘labelled’ as such, or may only be provided to people entering a bariatric surgery pathway.(6, 7) Thus, while the National Institute for Care and Health Excellence (NICE) has published 23 ‘products’ relating to obesity including a guideline on ‘Obesity: identification, assessment and management’,(8) their evaluations and recommendations are not explicitly linked to the tiered service approach to care.

When the tiered obesity services were first introduced it was envisaged the then Clinical Commissioning Groups took responsibility for Tier 3 and Tier 4 services. (9) Following the passage of the 2022 Health and Care Act, Integrated Care Systems (ICS) were formalised as legal entities with statutory powers and responsibilities. The ICSs comprise **integrated care boards (ICBs)** responsible for planning and funding most NHS services in the area and **integrated care partnerships (ICPs),** statutory committees that aim to tackle and improve inequalities in access and outcomes in population health care. (10, 11)

Access to specialist weight management services is known to be patchy across England. In 2022, the Royal College of Physicians (RCP) reported that a third of the population had no access to these services and, where they did exist, waiting times were unacceptable.(9) They went on to suggest that Integrated Care Systems (ICS) would be well-placed to co-ordinate the identification and referral of patients from primary to secondary or tertiary care(12) and that every ICS should have a weight management centre able to support and treat children, young people and adults. There is known to be a two-way disconnect between commissioning and provision of Tier 3 services. In order to obtain better information on service provision, an NHS national audit of Tier 3 service providers in England was launched in April 2022. In March 2023 (the latest data available) only 15 providers had entered data; they reported 1500 referrals ‘with at least one BMI record’. (13)

A recent cohort study accessed routinely collected primary care data in England from the Clinical Practice Research Datalink linked with Hospital Episode Statistics on just under 2 million adults. (14) They found that, despite national guidance, between 2007 and 2020, of 246,496 individuals with a BMI ≥ 35.0, only 16% received a weight management (WM) referral, and 3,701 (1.09% of those with severe and complex obesity) underwent bariatric surgery. They concluded that ‘that access to WM programmes in England is very low and has not improved over the last 10 years. They also highlighted regional inequalities in access to WM interventions across England.

In October 2023 and March 2024, NICE published early value assessments of digital technologies for delivering multi-disciplinary weight-management services (15) and approved seven digital services for both prescribing and monitoring weight-management medicine and delivering multidisciplinary weight-management services. These value assessments post-dated our survey dates and were not aligned to the definition of Tier 3 services.

The past decade has seen a step-change in the efficacy of medical treatments for obesity (16) with intensive meal-replacement diets and the licensing of anti-obesity medicines (AOM) capable not only of producing clinically significant weight loss, but also reversing type 2 diabetes, and reducing cardiovascular morbidity and mortality. (17, 18). AOMs were envisaged to be included within Tier 2 services, but in reality, they are almost exclusively available through Tier 3 services. With this background we sought to evaluate Tier 3 service commissioning in England.

### Aim

The aim of this study was to map, evaluate the content and scale of current commissioning of Tier 3 services by integrated care boards (ICB)s in England and explore how well they meet their population needs.

## Methods

In September 2023 we sent a freedom of information (FOI) request to all ICSs in England requesting the following information:

1. Details of Tier 3 Pathways:

- Please provide a comprehensive outline of the Tier 3 obesity care pathways within your ICB.
2. Providers of Tier 3 Pathways:

- Include the names of providers involved in delivering Tier 3 services within your ICB during the timeframe April 1, 2022, and March 31, 2023.
3. Volume of Referrals

- Please disclose the total number of referrals made to Tier 3 obesity care services within your ICB during the same period of time (April 1 2022, and March 31 2023).
4. Patients Commencing Tier 3 Treatment

- Provide the number of patients who commenced Tier 3 obesity treatment within your ICB during the same period (April 1 2022 to March 31, 2023).

### Data handling and analysis

Where data were provided, we collated and grouped service provision by whether it was NHS, non-NHS, a combination of the two or not provided. We further evaluated the services provided as to whether they offered: face-to-face care only (F2F): online only: a combination of the two: or not provided. We also recorded whether numbers of referrals into Tier 3 services were collected and, if so, the number of referrals and individuals starting treatment between April 12022 and 31 March 2023.

Neither data on obesity prevalence in adults at ICS/ICB level, nor UK prevalence of BMI>35kg/m2 are available.(personal communication Office for Health Improvement and Disparities, Department of Health, and Social Care), We therefore estimated the number of people eligible for Tier 3 services at a regional level through two scenarios: scenario 1 assumed that 10% of the adult population had obesity class II and III (1); scenario 2 used Canadian(19) and Australian data(20)) that showed 28% of people with obesity have a Body Mass Index (BMI) ≥35 kg/m2. Deprivation is the major underlying inequality underpinning differences in obesity levels (21, 22), so we explored the relationship with deprivation measured by the Index of Multiple Deprivation IMD (23) for each ICB and referral rates to Tier 3 services (Pearson Correlation, Microsoft Excel for Mac, 16.86). Results

We received responses from all 42 ICSs, a 100% response rate. Five (12%) ICSs reported that their ICB had no Tier 3 provision (NHS North West London, North Central London, Lancashire and South Cumbria, Northeast and North Cumbria, and Northamptonshire.) Table 1 shows the type of provision in terms of NHS, Non-NHS or a combination of the two. Exclusively NHS services were provided by 26 (62%) ICSs. Nearly two-thirds of ICBs offering treatment or providing data offered a combination of face-to-face and online appointments, however, 10 (24%) either did not respond, did not provide services, or did not know what they had commissioned (Table 1). The geographical distributions of these responses are shown in Figure 1. Twenty-four (57%) of ICBs collected data on the number of referrals made within their commissioned services;15 reported that data were not available, and 3 did not know. ICBs across the Midlands were the least likely to have these data. (Figure 2)

**Table 1.** Type of provision of Tier 3 services.

| Provision Type | Count | Percentage | Method | Count | Percentage |
| --- | --- | --- | --- | --- | --- |
| Blended | 8 | 19% | Blended (Face-to-face and online) | 25 | 60% |
| NHS provision | 26 | 62% | Face-to-face only | 2 | 5% |
| Non-NHS provision | 3 | 7% | Online only | 1 | 2% |
| No provision | 5 | 12% | No provision | 4 | 10% |
|  |  |  | No response/unknown | 10 | 25% |
| **TOTAL** | **42** |  | **TOTAL** | **42** |  |

**Figure 1:**
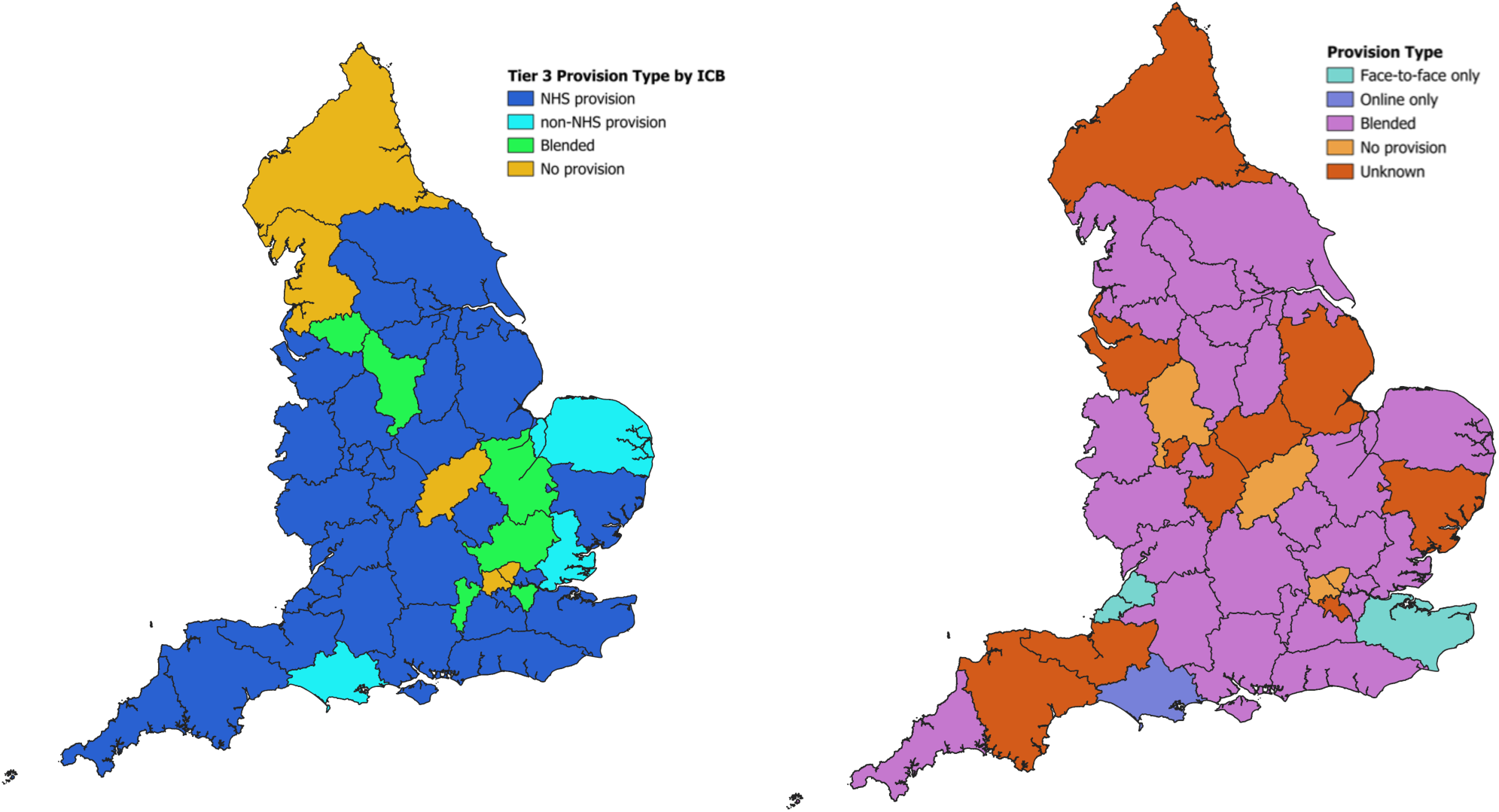
Geographical distribution of type of provision of Tier 3 management (left) and method of service delivery (right) Map showing geographical distribution of type of Tier 3 services: mainly NHS provided, but areas with non-provision in orange, non-NHS in light blue, and blended in green. Map showing geographical distribution of type of Tier 3 services: a small minority face-to-face or online only in dark and light blue, but most blended in purple.

**Figure 2.**
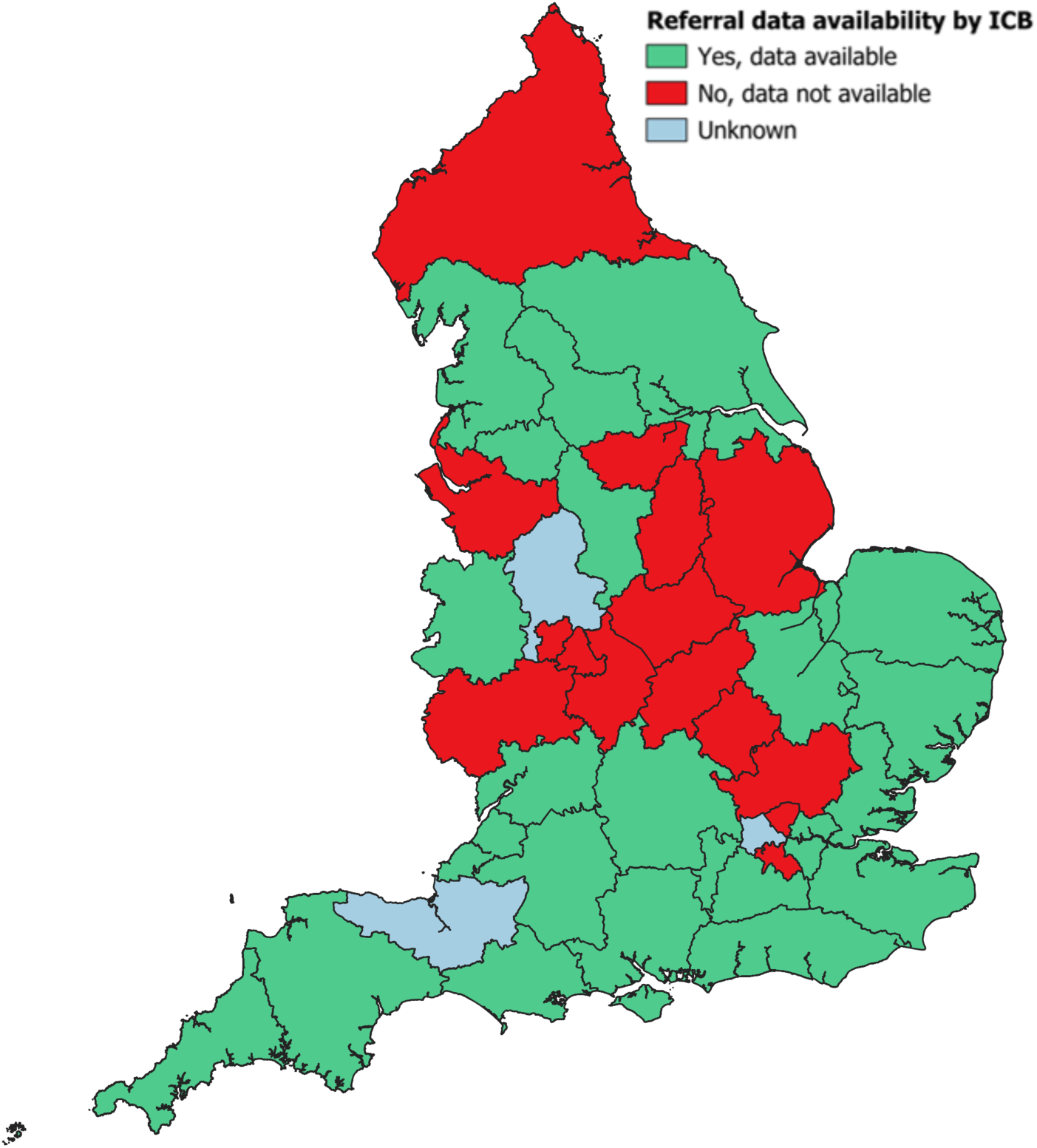
Referral data availability by ICB. Map to show distribution by Integrated Care Board of those with referral data available (green), unavailable (red) or unknown (light blue). Concentration of unknown data from North of England and Midlands.

Table 2 shows the number of Tier 3 referrals and the number of patients starting treatment in the year April 2022 to March 2023 by ICB. Figure 3 shows the large variation in access to treatment with the Midlands and North East and South East most poorly served.

**Table 2.** Number of referrals to Tier 3 and number starting treatment, April 2022-March 2023.

| NHS ICB | Referrals | Treatment |
| --- | --- | --- |
| NHS Greater Manchester ICB | 12,416 | 2145 |
| NHS South East London ICB | 3,397 | 1711 |
| NHS Devon ICB | 2,732 |  |
| NHS Sussex ICB | 2,658 | 2321 |
| NHS Mid and South Essex ICB | 2,605 | 630 |
| NHS Norfolk and Waveney ICB | 2,400 | 486 |
| NHS Humber and North Yorkshire ICB | 2,269 |  |
| NHS Hampshire and Isle of Wight ICB | 2,105 | 1163 |
| NHS Lancashire and South Cumbria ICB | 2,105 | 1163 |
| NHS West Yorkshire ICB | 1,790 |  |
| NHS Gloucestershire ICB | 1,406 | 67 |
| NHS Derby and Derbyshire ICB | 1,400 | 296 |
| NHS Kent and Medway ICB | 1,163 | 1111 |
| NHS Cambridgeshire and Peterborough ICB | 939 | 544 |
| NHS Bristol, North Somerset and South Gloucestershire ICB | 929 | 518 |
| NHS Cornwall and the Isles of Scilly ICB | 684 |  |
| NHS Shropshire, Telford and Wrekin ICB | 536 | 470 |
| NHS Surrey Heartlands ICB | 506 | 24 |
| NHS Dorset ICB | 503 | 234 |
| NHS Bath & North East Somerset, Swindon & Wiltshire ICB | 500 |  |
| NHS Buckinghamshire, Oxfordshire and Berkshire West ICB | 468 | 118 |
| NHS Frimley ICB | 337 | 207 |
| NHS Suffolk and North East Essex ICB | 314 | 173 |
| NHS North East London ICB | 229 | 124 |
| NHS Bedfordshire, Luton and Milton Keynes ICB |  | 603 |
| NHS Hertfordshire and West Essex |  | 274 |

**Figure 3.**
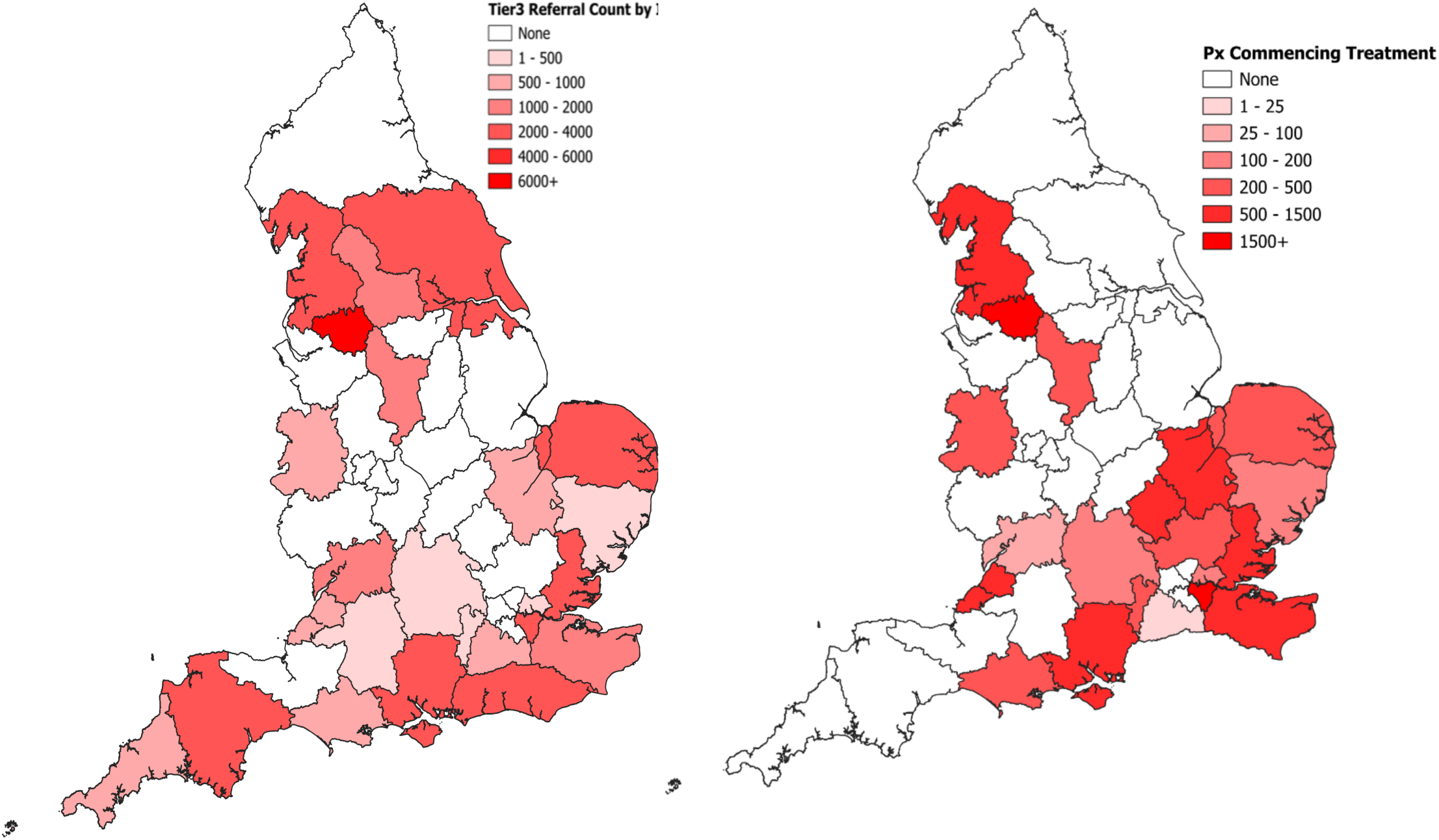
Referrals and treatment by geographical distribution. distribution of volume of Tier 3 services; no referrals in white and increasing numbers (maximum band >6000) by increasing shade of red. Highest areas of ‘none’ in Midlands and North England. Map showing geographical distribution numbers of patients starting Tier 3 services; none in white and increasing numbers (maximum band >1500) by increasing shade of red. Highest numbers of patients starting treatment in South East and North West England.

Figure 4 show the relationship between IMD for each ICB and referral rates to Tier 3 services. Including only data from those ICBs returning numbers of referrals identified, there was no correlation (Figure 5) between IMD and referral rates (R = 0.074; P=0.210).

**Figure 4.**
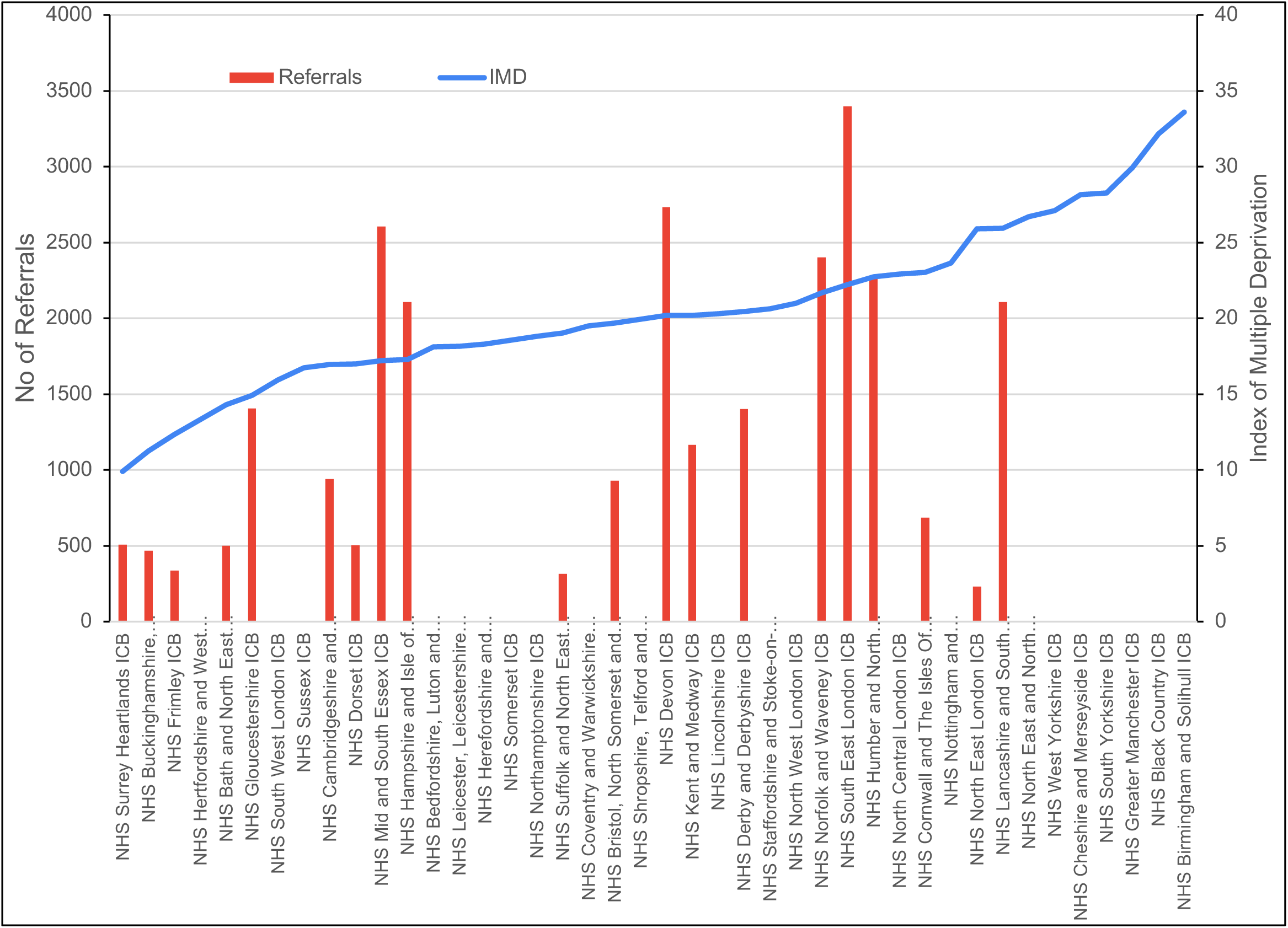
Number of referrals (vertical bars) and Index of Multiple Deprivation by ICB. Bar graph demonstrating lack of relation between referrals to Tier 3 (red bars) and increasing Index of Multiple Deprivation across Integrated Care Boards (blue line)

**Figure 5.**
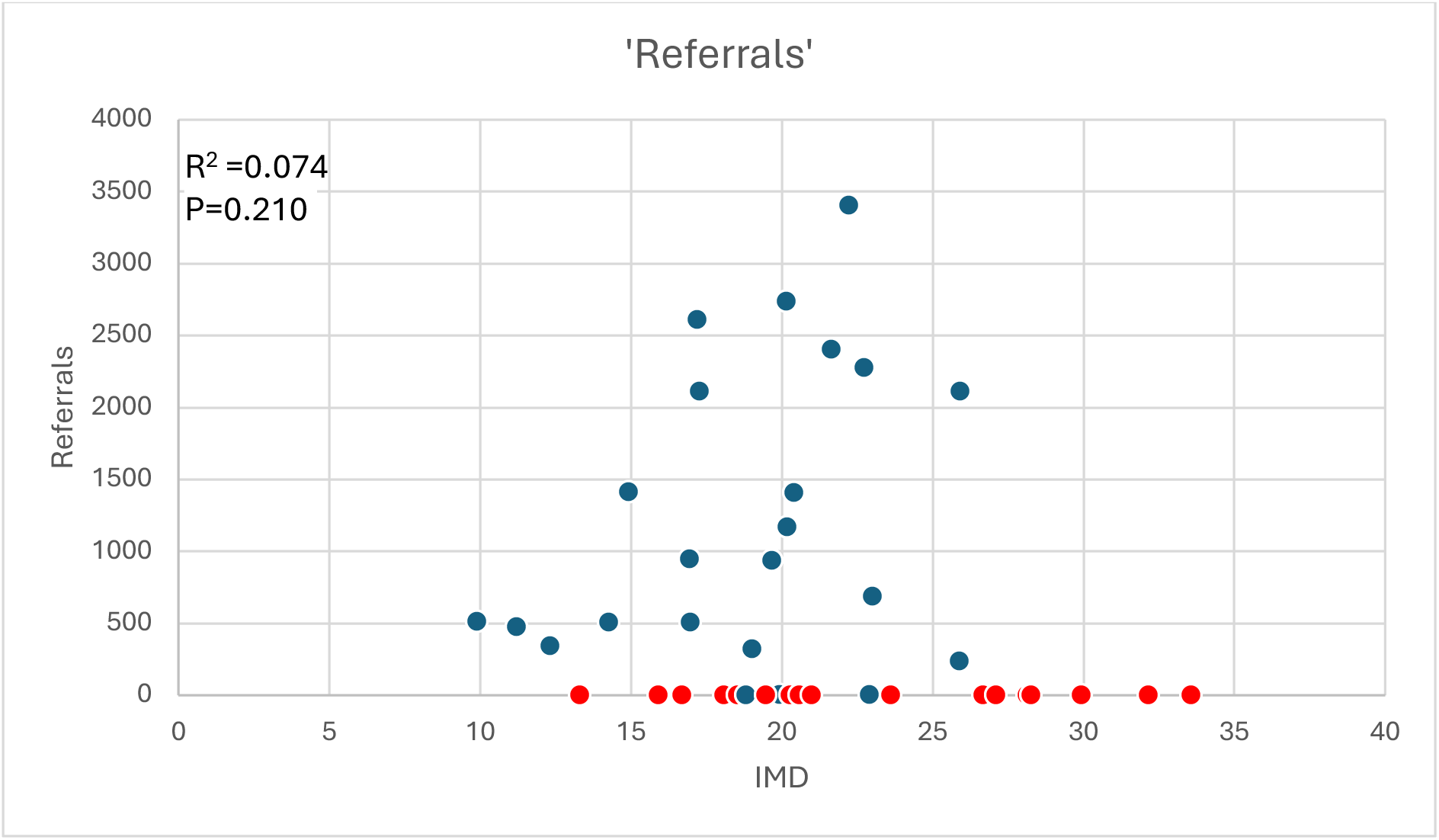
Relationship of Index of Multiple Deprivation score to number of referrals. Note where ICB returned that they had no data, the points are coloured red. Note: Linear regression excluded ICBs without data. Graph showing lack of correlation between Index of Multiple Deprivation score (x-axis) with referral numbers (y-axis) as blue dots. Red dots represent areas with no data available.

Since obesity prevalence data mapped to ICB level were not available, we used regional data and estimated that 25% of people with obesity were likely to have a BMI≥35 and be eligible for referral to Tier 3 management. We then calculated the percentage receiving treatment (Table 3). According to ICB reports the percentage of eligible patients treated ranged from 0 to 1.1%, with the highest number located in the South East of England, a region with the lowest levels of deprivation.

**Table 3.** Percentage of those eligible, receiving Tier 3 treatment.

|  | No starting Treatment | Obesity Prevalence | IMD | Population | Regional Population size | BMI>35 | % Treated |
| --- | --- | --- | --- | --- | --- | --- | --- |
| East of England |  | 27.3 |  |  | 5470092.0 | 373,334 | 0.73 |
| NHS Bedfordshire, Luton and Milton Keynes ICB | 603 |  | 18.11 | 1,070,212.00 |  |  |  |
| NHS Cambridgeshire and Peterborough ICB | 544 |  | 16.97 | 1,008,472.00 |  |  |  |
| NHS Hertfordshire and West Essex ICB | 274 |  | 13.32 | 1,612,064 |  |  |  |
| NHS Mid and South Essex ICB | 630 |  | 17.22 | 1,256,523.00 |  |  |  |
| NHS Norfolk and Waveney ICB | 486 |  | 21.67 | 1,086,462.00 |  |  |  |
| NHS Suffolk and North East Essex ICB | 173 |  | 19.04 | 1,048,423.00 |  |  |  |
| London |  | 27 |  |  | 7,118,942 | 480,529 | 0.38 |
| NHS North Central London ICB | 0 |  | 22.92 | 1,734,061 |  |  |  |
| NHS North East London ICB | 124 |  | 25.9 | 2,342,205.00 |  |  |  |
| NHS North West London ICB |  |  | 21 | 2,725,166.00 |  |  |  |
| NHS South East London ICB | 1711 |  | 22.24 | 2,051,571.00 |  |  |  |
| NHS South West London ICB |  |  | 15.93 | 1,726,507 |  |  |  |
| Midlands |  | 27.2 |  |  | 11,578,125 | 787,313 | 0.04 |
| NHS Birmingham and Solihull ICB |  |  | 33.59 | 1,577,949.00 |  |  |  |
| NHS Black Country ICB |  |  | 32.16 | 1,277,444.00 |  |  |  |
| NHS Coventry and Warwickshire ICB |  |  | 19.49 | 1,052,979.00 |  |  |  |
| NHS Derby and Derbyshire ICB | 296 |  | 20.43 | 1,111,009.00 |  |  |  |
| NHS Herefordshire and Worcestershire ICB |  |  | 18.28 | 818,249.00 |  |  |  |
| NHS Leicester, Leicestershire and Rutland ICB |  |  | 18.16 | 1,185,265.00 |  |  |  |
| NHS Lincolnshire ICB |  |  | 20.29 | 806,534.00 |  |  |  |
| NHS Northamptonshire ICB | 0 |  | 18.82 | 814,554.00 |  |  |  |
| NHS Nottingham and Nottinghamshire ICB |  |  | 23.64 | 1,240,698.00 |  |  |  |
| NHS Shropshire, Telford and Wrekin ICB | 0 |  | 19.95 | 521,391.00 |  |  |  |
| NHS Staffordshire and Stoke-on-Trent ICB |  |  | 20.61 | 1,172,053.00 |  |  |  |
| North East and Yorkshire |  | 27.9 |  |  | 9,012,300 | 628,608 | 0.00 |
| NHS Humber and North Yorkshire ICB |  |  | 22.75 | 1,771,076.00 |  |  |  |
| NHS North East and North Cumbria ICB |  |  | 26.7 | 3,139,823.00 |  |  |  |
| NHS South Yorkshire ICB |  |  | 28.27 | 1,483,968.00 |  |  |  |
| NHS West Yorkshire ICB |  |  | 27.11 | 2,617,433.00 |  |  |  |
| North West |  | 27.8 |  |  | 4,956,954 | 344,508 | 0.34 |
| NHS Cheshire and Merseyside ICB |  |  | 28.16 | 2,714,167 |  |  |  |
| NHS Greater Manchester ICB |  |  | 29.94 | 3,146,943.00 |  |  |  |
| NHS Lancashire and South Cumbria ICB | 1163 |  | 25.92 | 1,810,011.00 |  |  |  |
| South East |  | 27.1 |  |  | 6,625,901 | 448,905 | 1.10 |
| NHS Buckinghamshire, Oxfordshire and Berkshire West ICB | 118 |  | 11.23 | 1,935,027.00 |  |  |  |
| NHS Frimley ICB | 207 |  | 12.34 | 808,083.00 |  |  |  |
| NHS Hampshire and Isle of Wight ICB | 1163 |  | 17.28 | 1,916,638.00 |  |  |  |
| NHS Kent and Medway ICB | 1111 |  | 20.19 | 1,966,153.00 |  |  |  |
| NHS Surrey Heartlands ICB |  |  | 9.9 | 1,122,802 |  |  |  |
| NHS Sussex ICB | 2321 |  | 16.72 | 1,820,464 |  |  |  |
| South West |  | 27.1 |  |  | 5,329,585 | 361,079 | 0.20 |
| NHS Bath and North East Somerset, Swindon and Wiltshire ICB |  |  | 14.29 | 980,516.00 |  |  |  |
| NHS Bristol, North Somerset and South Gloucestershire ICB | 518 |  | 19.69 | 1,057,832.00 |  |  |  |
| NHS Cornwall and The Isles of Scilly ICB |  |  | 23.03 | 601,786.00 |  |  |  |
| NHS Devon ICB |  |  | 20.17 | 1,273,431.00 |  |  |  |
| NHS Dorset ICB |  |  | 16.99 | 819,184.00 |  |  |  |
| NHS Gloucestershire ICB | 199 |  | 14.93 | 1,820,464 |  |  |  |
| NHS Somerset ICB |  |  | 18.55 | 596,836.00 |  |  |  |

**Table 4.** Non-NHS Provision of Tier 3 services.

| **Name of Service** | **Multidisciplinary Approach including physician** | **Comprehensive Assessment including face-to-face consultation and screening** | **Personalized Treatment Plans including pharmacotherapy** | **Ongoing Support and Monitoring** | **Published Evaluation and Quality Improvement** |
| --- | --- | --- | --- | --- | --- |
| Everyonehealth (29)<br>HealthyYou (30) | No | No | No | Yes | Not stated |
| Morelife (31) | Yes | No | Yes | Yes | No |
| Oviva (32) | No | No | No | Yes | Yes |

## Discussion

This comprehensive survey of the commissioning of Tier 3 specialised obesity services in England, accessed through freedom of information requests to Integrated Care Systems, demonstrates the suboptimal of provision of care and treatment to people living with obesity. Less than 1% of people with a BMI ≥35 kg/m^2^ received treatment between March 2022 and April 2023. These findings accord with an earlier Clinical Practice Research Datalink (CPRD) study that found that just 3% of those with a recorded diagnosis of overweight/obesity in England between 2007 and 2020 had a referral for weight management recorded (14). Given that the 31% of adults with recorded overweight and obesity in CPRD represents about half the estimate of 64% by the Health Survey for England, the authors concluded their findings would translate to only 1.5% of the truly eligible population receiving a weight management referral. Additionally, there was considerable variation in the provision of services that did not relate to local needs as assessed by obesity prevalence or deprivation, a major determinant of obesity (22). While the period covered by our survey was soon after the establishment of ICBs, there has been little, if any, improvement since reports in 2013(24), and 2016(9).

Five of the 42 areas in England have no commissioned Tier 3 obesity services. Of equal concern is that even where Tier 3 services are commissioned, many do not meet NICE or other Commissioning guidelines. This results in an inability of people living with obesity to access the growing availability of both medical and surgical treatments for obesity with proven safety and efficacy. Based on the returns we received it is likely that few, if any, of the ICSs would be able to audit the outcomes and efficacy of their commissioned services. However, exclusively online treatment programmes are currently: not complete in the composition of their multidisciplinary team; without clinical assessment and screening for obesity related complications; without access to pharmacotherapy or surgery and do not comply with NICE guidance.

Due to the high prevalence of people eligible for medical (e.g. pharmacological) and/or surgical treatments, ICBs may need initially to prioritise who should receive treatment. This would need to be based upon guidance from the medical profession as well as patients living with obesity. In February 2024 the government stated ‘We are committed to the safe introduction of new weight loss drugs into the NHS and are actively exploring ways to increase access to more people who meet the relevant eligibility criteria’ (25). Well placed to drive the development and provision of obesity services are: the chair of the Obesity Mission (identified as one of the key healthcare missions in the 2021 <u>Life Sciences Vision</u> focussing on how life sciences interventions can be used to treat obesity) (26) and the newly appointed NHS England National Clinical Director for diabetes and obesity. We recommend that ICBs urgently review the adequacy of their Tier 3 service commissioning, both in terms of quantity and quality and make it a priority to address the current treatment gap. The need for this is highlighted by the recent report that one in six ICBs in England have stopped accepting new patients for specialist weight management services. (27) It is imperative that we do not neglect expansion of non-surgical obesity services, paying particular attention to areas with little or no resource. (28)

## Conclusions

Provision of Tier 3 services in England is inadequate and variable, they currently fail to meet the needs of the population, with 5 regions providing no Tier 3 services at all. Areas with highest levels of deprivation provided the most limited access. Even where commissioned, services often do not meet commissioning guidelines. Action is urgently needed to implement Health Service policy and ensure services conform with clinical need and national guidelines.

## Authors’ contribution

Conceptualisation: Nick Finer, Cecilia Pyper and Nikolaos Fragkas

Results and analyses: Nick Finer, Cecilia Pyper

Writing original draft: Nick Finer, Cecilia Pyper

Review and editing: Nick Finer, Cecilia Pyper, Nikolaos Fragkas, Alexander Dimitri Miras, Dimitri J Pournaras, Sarah Le Brocq and John Wass.

## Patient and public involvement

Co-author (SLB) as director and founder of the CIC All About Obesity represents patients’ perspectives.

## Ethics approval

Not applicable

## Data availability statement

All data relevant to the study are included in the article or uploaded as supplementary information.

